# Left and Right Ventricular Diameters Independently Predict Outcome in Intermediate and High-Risk Pulmonary Embolisms

**DOI:** 10.1101/2024.05.16.24307515

**Authors:** Daniel Spevack, Atul D. Bali, Arjun Kanwal, Ameesh Isath, Tanya Sharma, Syed Ahsan, Ramin Malekan, Joshua B. Goldberg

## Abstract

**Background:** The right to left ventricular diameter ratio (RV/LV) is a key imaging parameter used in risk stratification of pulmonary embolism. While having RV/LV above a cutoff value of 0.9 is usually attributed to pathological RV dilatation, we hypothesized that increased RV/LV may also be attributable to reduced LV filling. We aimed to examine RV and LV diameters as independent predictors of PE outcomes in patients presenting with intermediate or high-risk PE.

**Methods:** We measured RV and LV diameters on pre-intervention echocardiograms performed on 164 subjects presenting at or referred to our center for large PE requiring surgical thrombectomy, venoarterial extracorporeal membrane oxygenation (VA-ECMO), or catheter directed thrombectomy.

**Results:** The primary outcome of death (n=3) or survival of cardiopulmonary resuscitation (CPR) (n=11) was met in 14 subjects. Smaller LV diameter and larger RV diameter were each demonstrated to be independent predictors of the primary outcome. LV diameter of <3.6 cm, RV diameter >5.0 cm, and RV/LV ratio of >1.2 were the cutoff values for each parameter most associated with the primary outcome.

**Conclusion:** This study demonstrates that, in addition to dilated RV, small LV is an independent risk factor of death or need for CPR in hemodynamically significant PE treated with advanced therapies. This finding suggests that the predictive value of RV/LV for PE outcomes may be attributed to both RV enlargement and reduced LV filling.

## Introduction

Acute pulmonary embolism (PE) is the third leading cause of cardiovascular mortality behind myocardial infarction and stroke (1). The majority of PE associated deaths are attributed to acute right ventricular (RV) dysfunction. Accordingly, PE patients who present with significant RV dysfunction have a high associated PE mortality ranging between 25 to 65% in high-risk PE with little improvement in mortality over the last 2 decades. (2,3)

PE associated RV dysfunction is caused by an abrupt increase in RV afterload with ensuant pressure-volume overload. Numerous, adverse structural and functional changes occur including RV dilation, RV subendocardial ischemia, and increased pulmonary vascular resistance. (10) RV dilation and increased pulmonary resistance both adversely affect LV filling due to decreased transpulmonary blood flow and bowing of the interventricular septum toward the left ventricle. (10) Decreased LV preload is compensated with increased adrenergic tone manifest as tachycardia and increased systemic vascular resistance. Hemodynamic decompensation occurs when compensatory mechanisms fail and the LV is no longer able to support systemic perfusion.

Structural assessment of the RV is integral to risk stratification of PE patients and decision to pursue advanced interventions. The ratio of the RV and LV diameter (RV/LV) is a core component of current PE risk stratification models with RV/LV> 0.9 used as a cut off for pathological RV dilation in the setting of PE in all major guidelines and trials. (4,5-8,11) Since underfilling of the LV is also a key feature of severe PE, we hypothesized that both increased RV diameter and decreased LV diameter would be independent predictors of PE prognosis in those presenting with hemodynamically significant PE.

## Methods

Patients presenting between 2006 and 2022 with PE and RV dysfunction treated with surgical embolectomy, VA-ECMO, and/or transcatheter suction embolectomy who had adequate TTE images acquired prior to intervention were prospectively collected in a single institution clinical database. Acute PE were diagnosed with Computer Aided Tomographic Angiography (CTA) in the entire study population. All patients met diagnostic criteria for either intermediate or high-risk PE as defined by the European Society of Cardiology criteria. (11) High-Risk PE was defined as acute PE with RV dysfunction and hemodynamic instability. Intermediate risk PE was defined as hemodynamically stable acute PE with RV dysfunction. RV dysfunction was defined as an RV/LV ratio > 0.9 (as measured on CT angiogram or TTE) and/or elevated troponin-I >0.03 ng/mL and/or brain natriuretic peptide (BNP) >100 pg/mL. Hemodynamic instability was defined as a systolic blood pressure less than 90 mmHg for greater than 15 minutes, requirement for inotropes or pressors, cardiac arrest or drop in systolic blood pressure more than 40 mmHg from one’s baseline pressure. The Institutional Review Board of New York Medical College/Westchester Medical Center approved this study under the “consent exempt” category.

### RV dysfunction assessment

Pre-intervention transthoracic echocardiograms were reviewed using IntelliSpace Cardiovascular release 4.2 software (Phillips Medical Systems, Andover, MA). Measurements were performed by one investigator (SA) for studies performed between 2006 and 2019 and subsequently by a separate investigator (AK) for studies performed from 2020 to 2021. In the apical 4-chamber view, LV and RV diastolic diameters were measured at the level of the mitral and tricuspid valve tips at end diastole, Figure 1. The LV and RV were generally measured on the same end diastolic frame, depending on the endocardial image quality for each chamber. RV systolic and diastolic areas were also measured in the apical 4-chamber view on a RV focused view.

**Figure 1:**
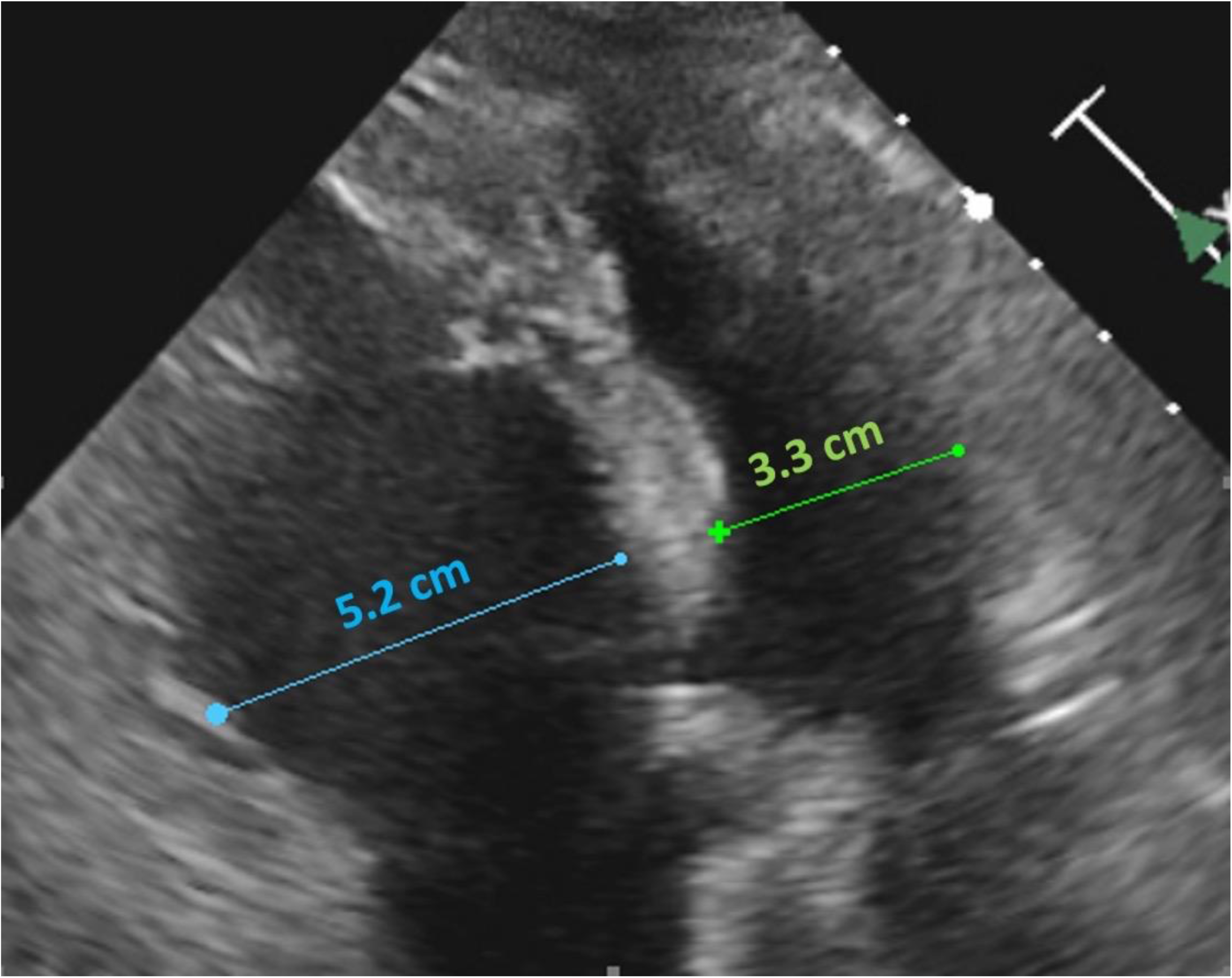
Representative echocardiogram in a subject with increased RV diameter and decreased LV diameter. Measurements were made on the apical 4-chamber view at the level of the mitral and tricuspid valve tips. Measurements were made at end-diastole.

**Figure 2:**
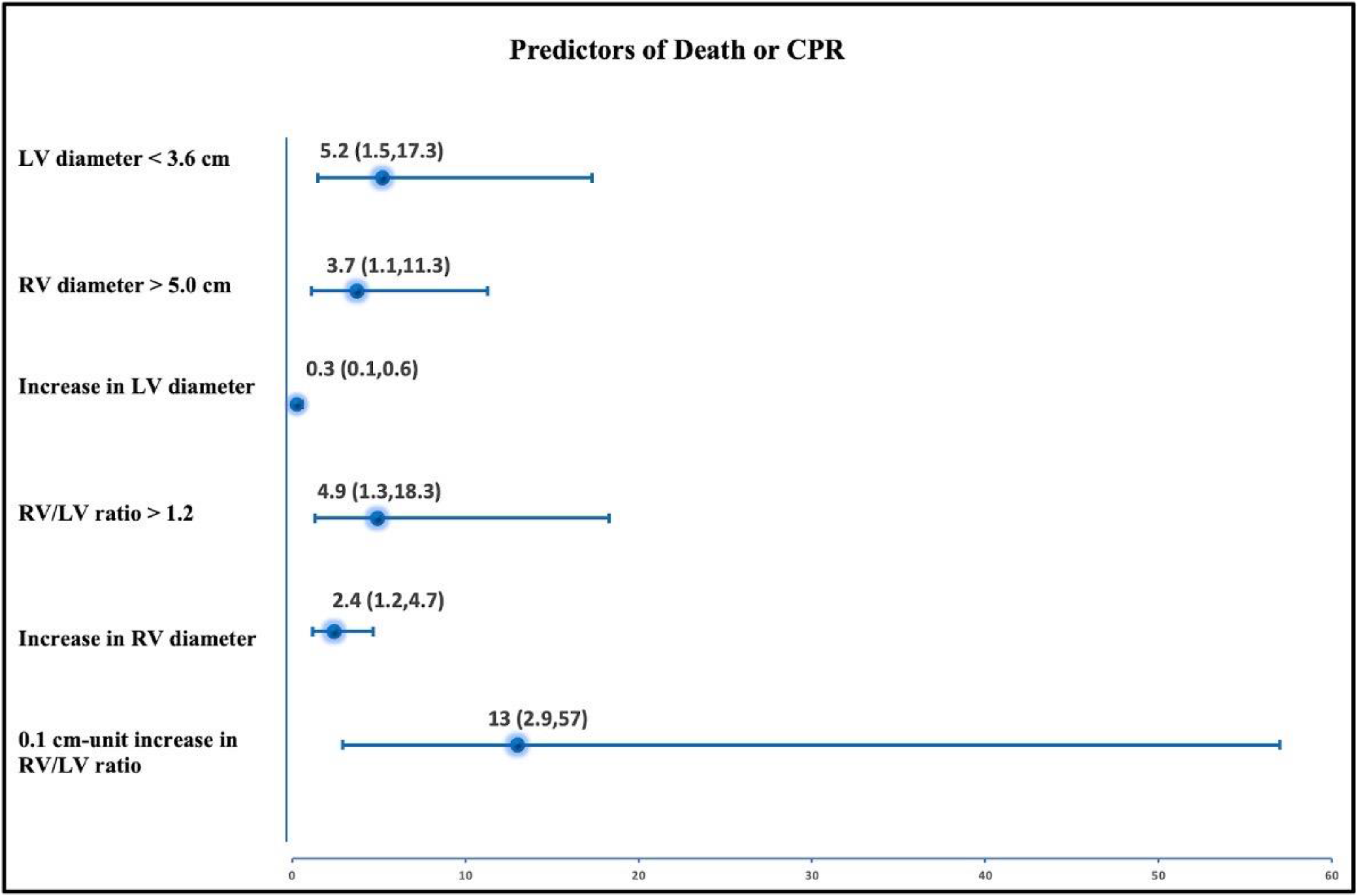
Forest plot of statistically significant echocardiographic parameters as predictors for death or requiring CPR. *LV* – left ventricle, *RV* – right ventricle, *CPR* – cardiopulmonary resuscitation.

### Inter-observer variability

To evaluate inter-observer variability, echocardiographic measurements of the 30 most recently enrolled subjects were independently reviewed by two investigators (AB and AK), both blinded to each other’s assessments. For each variable studied, we looked at the mean difference between the two reader’s “bias”. We also looked at “variability” between the reader measurements defined as the standard deviation of mean difference between readers.

### Outcomes

The primary outcome was the composite of death or survivors of cardiopulmonary arrest requiring cardiopulmonary resuscitation (CPR).

### Statistical analysis

Descriptive statistics were used to describe the demographic and clinical characteristics of the cohort. Categorical variables are presented as percentages and continuous variables as mean ± standard deviation (SD). Comparisons of continuous variables between groups were performed with t tests and of categorical variables by the Chi square test. A p-value less than 0.05 was considered significant. Statistical analysis was performed using Stata software, version 11.2 (College Station, TX).

## Results

Out of a total of 249 patients, 164 had adequate pre-procedure echocardiographic views to measure RV and LV diameters reliably. There were 14 subjects who experienced the primary outcome (3 deaths, 11 CPR survivors). The baseline demographics, comorbidities and presenting vital signs are displayed in the table stratified by the primary outcome, Table 1. Subjects who experienced the primary outcome more often had a history of cancer, presented with lower blood pressure and were more often classified as high risk. Otherwise, no significant differences were seen between the groups with respect to demographics, comorbidities, presenting symptoms and vital signs.

**Table 1:** Baseline demographics, comorbidities, presenting vital signs are stratified by the primary outcome.

|  | **Whole sample**<br>**(n=164)** | **Death or**<br>**CPR Survivor**<br>**(N=14)** | **Alive and**<br>**No CPR**<br>**(N=150)** | **p-**<br>**value** |
| --- | --- | --- | --- | --- |
| Age (years) | 59 ± 15 | 61 ± 13 | 58 ± 15 | 0.42 |
| Male sex (n ,%) | 95 (57.9%) | 9 (64.2%) | 86 (45.73%) | 0.66 |
| White (n ,%) | 111 (67.6%) | 10 (71.4%) | 101 (67.3%) | 0.42 |
| BMI (kg/m<sup>2</sup>) | 34.2 ± 8.1 | 35.8 ± 11.2 | 34 ± 7.8 | 0.44 |
| Recent surgery (n ,%) | 44 (267%) | 4 (28%) | 40 (26%) | 0.88 |
| Recent trauma (n ,%) | 11 (6.7%) | 1 (7.1%) | 10 (6.6%) | 0.97 |
| Prior thromboembolism (n ,%) | 31 (18.9%) | 1 (7.1%) | 30 (20%) | 0.21 |
| Cancer (n ,%) | 22 (13.4%) | 5 (35.7%) | 17 (11.3%) | 0.01 |
| COPD or asthma (n ,%) | 55 (33.5%) | 5 (35.7%) | 50 (33.3%) | 0.66 |
| Current or former smoker (n ,%) | 28 (17%) | 4 (28.5%) | 24 (16%) | 0.23 |
| Diabetes (n ,%) | 40 (24.3%) | 1 (7.1%) | 39 (26%) | 0.11 |
| Coronary artery disease (n ,%) | 13 (7.9%) | 2 (14.2%) | 11 (7.3%) | 0.36 |
| Congestive heart failure (n ,%) | 4 (2.4%) | 0 (0%) | 4 (2.6%) | 0.53 |
| Stroke (n ,%) | 11 (6.7%) | 1 (7.1%) | 10 (6.6%) | 0.95 |
| Systolic blood pressure (mmHg) | 112 | 89 ± 11.8 | 113 ± 28.6 | 0.008 |
| Heart rate (beats per minute) | 106 | 106 ± 19 | 106 ± 18 | 0.94 |
| Elevated biomarkers (n ,%) | 164 (100%) | 14 (100%) | 150 (100%) | 1.0 |
| Requiring pressors (n ,%) | 24 (14.6%) | 3 (21.4%) | 21 (14%) | 0.45 |
| High risk PE (n ,%) | 51 (31%) | 12 (85.7%) | 39 (26%) | < 0.001 |

### Echocardiographic Parameters

The baseline echocardiographic parameters and patient outcomes are displayed in the table stratified by the primary outcome, Table 2. All patients had RV dysfunction on presenting TTE. Overall, the mean RV/LV end-diastolic ratio was 1.2 ± 0.3 (range 0.7 to 2.8). Subjects who experienced the primary outcome had smaller measured LV diameter and increased RV:LV ratio, but did not have significantly larger RV diameter compared to those who did not. Higher RV/LV ratio used as a continuous variable was associated with increased odds of the primary outcome (odds ratio (OR) 13 [95% confidence interval (CI): 2.9, 57] per 1-unit increase, p=0.001. RV/LV ratio > 1.2 was the cutoff value most highly associated with the primary outcome, OR 4.9 [95% CI: 1.3, 18.3], p=0.02. Mean RVD and LVD on presentation were 4.5 ± 0.9 cm and 3.8 ± 0.8 cm respectively. As a continuous variable, increasing RVD (OR 2.4 [95% CI: 1.2, 4.7], p=0.01) and decreasing LVD (OR 0.3 [95%CI: 0.1, 0.6], p=0.001) per 1 cm change, were each independent predictors of death or CPR. Having RVD > 5.0 cm (OR 3.7 [95% CI: 1.2, 11.3], p 0.02) and LVD < 3.6 cm (OR 5.2 [95% CI: 1.5, 17.3], p=0.008) were the ventricle size cutoff values associated with the highest odds of the primary outcome. These cutoff values remained significant predictors even after adjustment for body surface area. Other parameters of RV size and function such as diastolic area, systolic area and fractional area change did not predict death or CPR.

**Table 2:** Baseline echocardiographic variables, initial interventional treatment strategy and patient outcomes are stratified by the primary outcome.

|  | **Whole sample**<br>**(n=164)** | **Death or**<br>**CPR Survivor**<br>**(N=14)** | **Alive and**<br>**No CPR**<br>**(N=150)** | **p-**<br>**value** |
| --- | --- | --- | --- | --- |
| RV diameter (cm) | 4.5 ± 0.9 | 4.8 ± 0.8 | 4.4 ± 0.8 | 0.10 |
| LV diameter (cm) | 3.8 ± 0.8 | 3.2 ± 0.7 | 3.8 ± 0.7 | 0.005 |
| RV:LV diameter ratio | 1.20 ± 0.32 | 1.5 ± 0.3 | 1.2 ± 0.3 | < 0.001 |
| Catheter embolectomy (n ,%) | 40 (24.3%) | 1 (7.1%) | 39 (26%) | 0.11 |
| Surgical embolectomy (n ,%) | 99 (60.4%) | 9 (64.2%) | 90 (60%) | 0.79 |
| ECMO (n ,%) | 24 (14.6%) | 4 (28.6%) | 20 (13.3%) | 0.12 |
| Death (n ,%) | 3 (1.8%) | 3 (21.4%) | 0 (0%) | < 0.001 |
| CPR survivors (n ,%) | 11 (6.7) | 11 (78.5%) | 0 (0%) | < 0.001 |

#### Interobserver measurement agreement

In general, the agreement between the readers was very good, reflecting in the bias and variability across several measurements. For the RVD, the between reader bias was -0.22 cm with a variability of ± 0.34 cm (p=0.002). For LVD, the between reader bias was 0.16 cm, with a variability of ± 0.34 cm (p=0.02). RV/LV ratio had a between reader bias of -0.1, with a variability of ± 0.12 (p<0.001). The RV area presented a bias of -0.88 cm^2^ with a variability of ± 1.7 cm^2^(p=0.009). Fractional area change (FAC) had a between reader bias of -1.63 and variability of ± 6.15 (p=0.16).

## Discussion

This single institution analysis of 164 acute PE patients with intermediate and high-risk PE treated with transcatheter embolectomy, surgical embolectomy or VA-ECMO demonstrated that LV and RV diameter measurements are independently associated with death or need for CPR. LV diameter of <3.6 cm and RV diameter >5.0 cm were the cutoff values for each parameter most associated with the primary outcome. In our patient sample, RV/LV > 1.2 was the cutoff value most associated with the primary outcome. It is well established that RV dysfunction is associated with adverse outcomes in patients with acute PE. (5-8) Our findings support these prior works by demonstrating that increased RV/LV ratio and increasing RV diameter are independently associated with death or CPR. The novel finding in this study is that decreased LV diameter is, at least, as important as RV diameter in predicting death or CPR.

The independent association of smaller LV diameter with the primary outcome is consistent with acute PE pathophysiology and may better explain and predict hemodynamic collapse than RV dilation alone. Along with RV dilation and dysfunction caused by volume pressure overload, the LV becomes underfilled secondary to decreased transpulmonary flow and ventricular interdependence with the bowing of the RV towards the LV resulting in a hyperdynamic, underfilled LV. Compensatory mechanisms such as increased adrenergic tone, ensuant tachycardia and vasoconstriction, preserve hemodynamics. Hemodynamic collapse and cardiac arrest occur when the compensatory mechanisms fail and the LV can no longer support systemic and myocardial perfusion. Accordingly, our study shows that a small LV is a potentially ominous sign.

The independent association of LV diameter on CPR or death potentially can assist in the prediction of patients who may decompensate. Currently there are few metrics that can accurately predict decompensation or death among acute PE patients. Metrics such as RV/LV ratio, cardiac biomarkers and hemodynamics have been used to risk stratify patients, but there are no currently identified metrics or tools to predict which “stable” patients are at risk of decompensation. Use of LV diameter may provide a useful indicator of increased risk PE patients regardless of their initial risk classification.

### Limitations

The present study was performed at a single institution collected in a selected cohort that required surgical or catheter-based interventions for PE. This may limit generalizability of our data to broader patient populations undergoing evaluation for PE. In our study, RV and LV diameters were measured on pre-intervention echocardiograms. We cannot comment on the use of these parameters from computed tomography images.

## Conclusion

This study demonstrates the novel finding that increased RV and decreased LV diameters were both strongly associated with short term risk of death or need for CPR. This finding suggests that the predictive value of RV/LV for short term PE outcomes may be attributed to a combination of both increased RV afterload and reduced LV preload.

## Data Availability

The authors confirm that the data supporting the findings of this study are available within the article [and/or] its supplementary materials.

---

**Central Illustration:**

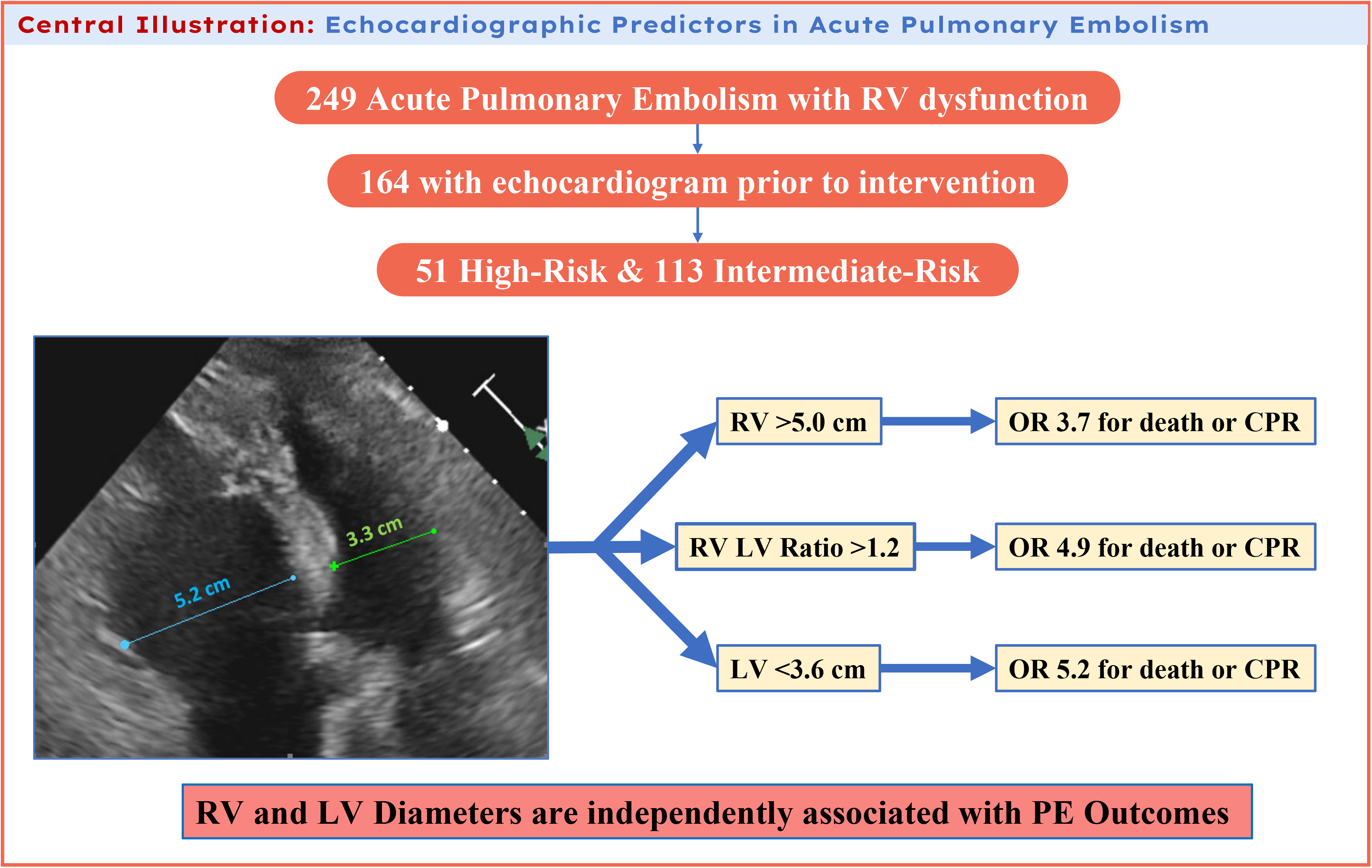

